# Sleep interventions for adults admitted to psychiatric inpatient settings: a systematic scoping review

**DOI:** 10.1101/2023.03.03.23286483

**Authors:** Anne M. Aboaja, Lindsay H. Dewa, Amanda E. Perry, Jon F. Carey, Rachel Steele, Ahmed Abdelsamie, Gies T. A. Alhasan, Ishwari S. Sharma, Scott A. Cairney

**Affiliations:** Forensic Service, Tees, Esk and Wear Valleys NHS Foundation Trust, UK; Mental Health and Addictions Research Group, University of York, York, UK; School of Public Health, Imperial College London, London, UK; Institute of Global Health Innovation, Imperial College London, London, UK; Library and Information Services, Tees, Esk and Wear Valleys, NHS Foundation Trust, UK; Department of Psychology, University of York, UK; York Biomedical Research Institute (YBRI), University of York, UK

**Keywords:** Psychiatric inpatients, Intervention, Sleep, Mental health, CBT for insomnia, Hospital

## Abstract

Sleep disturbances are common, affecting over half of adults with a mental disorder. For those admitted to a psychiatric ward, difficulties with sleep are compounded by factors relating to the inpatient setting. We conducted a scoping review of sleep intervention studies on adults admitted to psychiatric settings. We categorised the different types of sleep interventions and identified the effects on sleep and other health outcomes. Instruments used to measure sleep were also described. The search strategy yielded 2530 studies, of which 20 met the inclusion criteria. There was evidence of more non-pharmacological than pharmacological interventions having been tested in inpatient settings. Results indicated that non-pharmacological interventions based on cognitive behaviour therapy for insomnia improve sleep and may improve mental and physical health. Several distinct sleep measures were used in the studies. Objective sleep measures were not commonly used. Gaps in the literature were identified, highlighting the importance of research into a wider range of sleep interventions tested against a control using objective measures of sleep with evaluation of additional mental and physical health outcomes among adults in the psychiatric inpatient settings.

## Introduction

Good sleep is essential for our mental health, social functioning and quality of life. There is a complex bidirectional relationship between sleep and psychiatric disorders (1), many of which are associated with sleep continuity disturbances (2). Sleep disturbances commonly occur across mental disorders (3) and are risk factors for the onset (4) and prognosis of mental disorders (5). Sleep disturbances are present in up to 80% of people with psychosis (3, 6, 7) and up to 90% of people with depression (3, 7). Compared to people without a mental disorder, adults with schizophrenia have a shorter total sleep time (8), whilst those with bipolar disorder have a longer sleep duration (9). Both increased and reduced sleep duration is problematic. In addition, individuals with active symptoms of borderline personality disorder experience prolonged sleep onset latency (10). The relationship between sleep and mental disorders is further complicated by a third factor, physical health comorbidity, which is linked to both sleep and mental disorders. Compared to the general population, people with mental disorders have an increased risk of obesity, diabetes and cardiovascular disease (11), all of which are associated with sleep disturbances (12). In addition, the side effects of medication used to treat mental disorders not only directly affect sleep, but are also associated with an increased risk of developing these physical health conditions (13). However, targeting sleep disturbances directly through pharmacological and non-pharmacological interventions can improve sleep, and subsequently reduce these risks.

Maintaining a good sleep environment can also improve sleep. The psychiatric hospital is an inpatient setting which should be a therapeutic environment whereby improving mental and physical wellbeing is the goal. However, this is often not the reality. Features of the hospital environment can further disrupt sleep-wake regulation and further compound sleep disturbances (14) including inadequate daytime light exposure, noise at night (15), lack of autonomy (16, 17) and the ward regime (17). When compared to sleep at home, sleep in any hospital setting is significantly shorter in duration and poorer in quality (14), however, inpatient sleep quality is significantly poorer on psychiatric wards than non-psychiatric wards (18). Among adults with a wide range of mental disorders admitted to a psychiatric hospital, poor sleep quality, insomnia, and hypersomnia are common sleep problems (19). In contrast, sleep disorders of movement such as restless legs syndrome (16), sleep disorders of breathing such as sleep apnoea (16) and parasomnias are less prevalent (3).

Inadequate sleep negatively affects several areas of health including mood, cognition and behaviour (20, 21). Among people with mental disorders, sleep disturbances are associated with elevated levels of depression and irritability, as well as deficits in memory, concentration and decision-making (22). In the psychiatric inpatient setting, poor sleep is also linked to increased suicidality (23). Specifically in the secure psychiatric inpatient context, sleep disturbances have been associated with increased impulsivity (24) and aggression (25). The potential costs of untreated poor sleep among patients in a psychiatric hospital are significant. Patients who are unable to sleep during the night are more likely to sleep during the day and miss psychological therapies (26). Those with disturbed sleep who attend psychological therapies are more likely to experienced impaired learning due to poor attention (20) and memory (27, 28) and may be required to repeat inpatient therapies prior to discharge. Increased symptoms of mental disorder linked to poor sleep may result in a longer admission (29) and higher doses of psychotropic medication which has implications for physical health (30). Poor sleep impairs decision-making and increases impulsivity, suicidality and aggression (25, 31). Patients with impaired decision-making may experience reduced adherence to treatment. Those with increased suicidality and aggression carry a heightened risk of harm to their own lives and the lives of others and are likely to require a longer admission. Therefore, sleep improvement in the inpatient psychiatric setting is an opportunity to improve health outcomes and achieve financial benefits through the reductions in length of stay, psychotropic prescribing and missed or repeated psychological therapies.

The limited use of hypnotic medication when non-pharmacological sleep interventions are not beneficial is recommended for patients in psychiatric and non-psychiatric hospitals (32). Cognitive behavioural therapy for insomnia (CBT-i), melatonin supplements and, to a lesser degree, environmental modifications such as light therapy and aromatherapy, are the most effective non-pharmacological sleep interventions in non-psychiatric and psychiatric hospitals combined (33). Despite a growing body of literature on the use of sleep interventions in hospitals, the evidence specifically in psychiatric hospitals, where frequent patient observations are undertaken during the night (26), remains limited. Reviews have either excluded inpatients with a mental disorder (34), combined psychiatric populations with prison populations (35) or included them without separately examining their specific responses to sleep interventions (32, 33). Given the known associations between sleep, mental health and physical health, there is value in a review that systematically identifies studies of both pharmacological and non-pharmacological sleep interventions used among people with a mental disorder in psychiatric inpatient settings.

The aim of this scoping review was to (i) collate studies examining the effects of pharmacological and non-pharmacological sleep interventions specifically within the adult psychiatric inpatient setting, (ii) understand how sleep outcomes are measured in these studies and whether there are any barriers to measuring sleep and (iii) identify the effects these sleep interventions have on sleep and other health outcomes.

## Method

The protocol for this review was published (36). The review was undertaken in accordance with the Joanna Briggs Institute (JBI) scoping review methodological framework (37). We followed the Preferred reporting items for systematic reviews and meta-analyses (PRISMA) guidance to summarise the search results (38) and used the PRISMA extension for scoping reviews (PRISMA-ScR) to report other findings (39). The stages of JBI methodology included: (i) identification of research questions (ii) inclusion criteria (iii) search strategy (iv) study selection, (v) data extraction, (vi) presentation of results.

### Research questions

Review questions were:

1. how is sleep measured in sleep intervention studies undertaken in the adult psychiatric inpatient setting?
2. what barriers to measuring sleep are reported in sleep intervention studies in adult psychiatric inpatient settings?
3. which sleep interventions used in adult psychiatric inpatient settings have an effect on sleep and other health outcomes?

### Inclusion criteria

Quantitative studies were included if they examined the effectiveness of an intervention on sleep; a sleep parameter was a primary outcome; participants were adults aged 18 years and over; they were conducted in an inpatient psychiatric setting; and they were published in English. Study designs included randomised controlled trials, quasi-randomised trials and non-randomised/quasi-experimental studies. Mixed child and adult samples were excluded unless authors applied appropriate stratification by age in the data analysis, and therefore adults could be separated. Consistent with previous reviews, studies that focused solely on parasomnias, sleep apnoea, and sleep-related movement disorders were excluded (17, 25, 33, 34). Case studies and conference proceedings were also excluded.

### Search strategy

Four databases were searched: CINAHL, MEDLINE, PsycINFO and Web of Science. We identified studies using variations on the following concepts: “sleep” AND “mental” AND “hospital”. The detailed search strategy is outlined in the Supplement 1. Additional studies were identified through the first 1000 results on Google Scholar. No lower date limit was used. We conducted the search on 20 September 2019 and updated our search on 18 June 2020. We scanned the reference lists of included studies.

### Study selection

Identified articles were uploaded into EndNote version X9 (40) and duplicates were removed. One reviewer (AA1) conducted title and abstract screening. Two reviewers (AA2, GA) independently screened full text articles and where disagreement could not be resolved by discussion, resolution was achieved through discussion with a third reviewer (AA1).

### Data extraction

We used a pre-determined data extraction form which was modified while piloting data extraction from two identified studies. The modified data extraction form with additional items included: author, year of publication, country (additional item), study design, type of inpatient setting, intervention (name, format, facilitator, duration, frequency), intervention participants (gender, age, diagnosis), control, control participants (gender, age, diagnosis), sleep measure instrument/outcome (eg, nursing sleep chart/sleep duration), intervention effect, main sleep finding, barriers to measuring sleep sample characteristics, other health outcomes (emotional, cognitive, somatic) and study quality scores (additional item), as described below. Data extraction of all included studies was undertaken independently by two researchers, matching an academic (AP, LD, SC) with a clinician (AA2, GA, IS, JC) where possible to strengthen the academic rigour and clinical expertise applied during this phase. Reviewers within each pair then jointly checked data extraction content to ensure accuracy and resolved disagreements through discussion or, if required, with input from a third reviewer (AA1).

Whilst not a stipulated requirement for scoping reviews, we critically appraised all the included studies to understand the quality of the current literature. We used the Cochrane risk of bias tool (41) to assess the randomised controlled trials (RCTs). We also assessed the quality of all studies according to the five quality criteria of the Mixed methods appraisal tool (MMAT) (42). Using a recognised approach (43), the quality score for each study was expressed as a percentage (low: 0%, 20%; medium: 40%, 60%; high: 80%, 100%). The result allowed the overall quality of studies in the field to be estimated.

### Data presentation

Descriptive statistics were presented of study characteristics and samples. Sleep measures were categorised using a matrix of objective vs. subjective and validated vs. unvalidated and barriers to measuring sleep were noted. Studies were categorised according to intervention type: pharmacological or non-pharmacological. Non-pharmacological interventions were further divided into CBT-i based interventions and environmental interventions. Finally, we reported effects on sleep and other health outcomes according to intervention type. Where a measure of statistical significance was reported, sleep outcomes with effects sizes that were not statistically significant (i.e, effect sizes reported with a *p* value that was not less than 0.05) were excluded from tables or the text analysis. We synthesised the findings of the effect of interventions on sleep and other health outcomes by study design based on the presence of a comparison group.

### Patient and public involvement

The guidance for reporting involvement of patients and the public version 2 (GRIPP2) (44) was followed to report patient and public involvement (Supplement 2).

## Results

### Selection of studies

Figure 1 shows the results of the literature search and study selection in an adapted PRISMA flow diagram.(38) We identified 2530 unique studies and included 20 studies in the final review.

**Figure 1:**
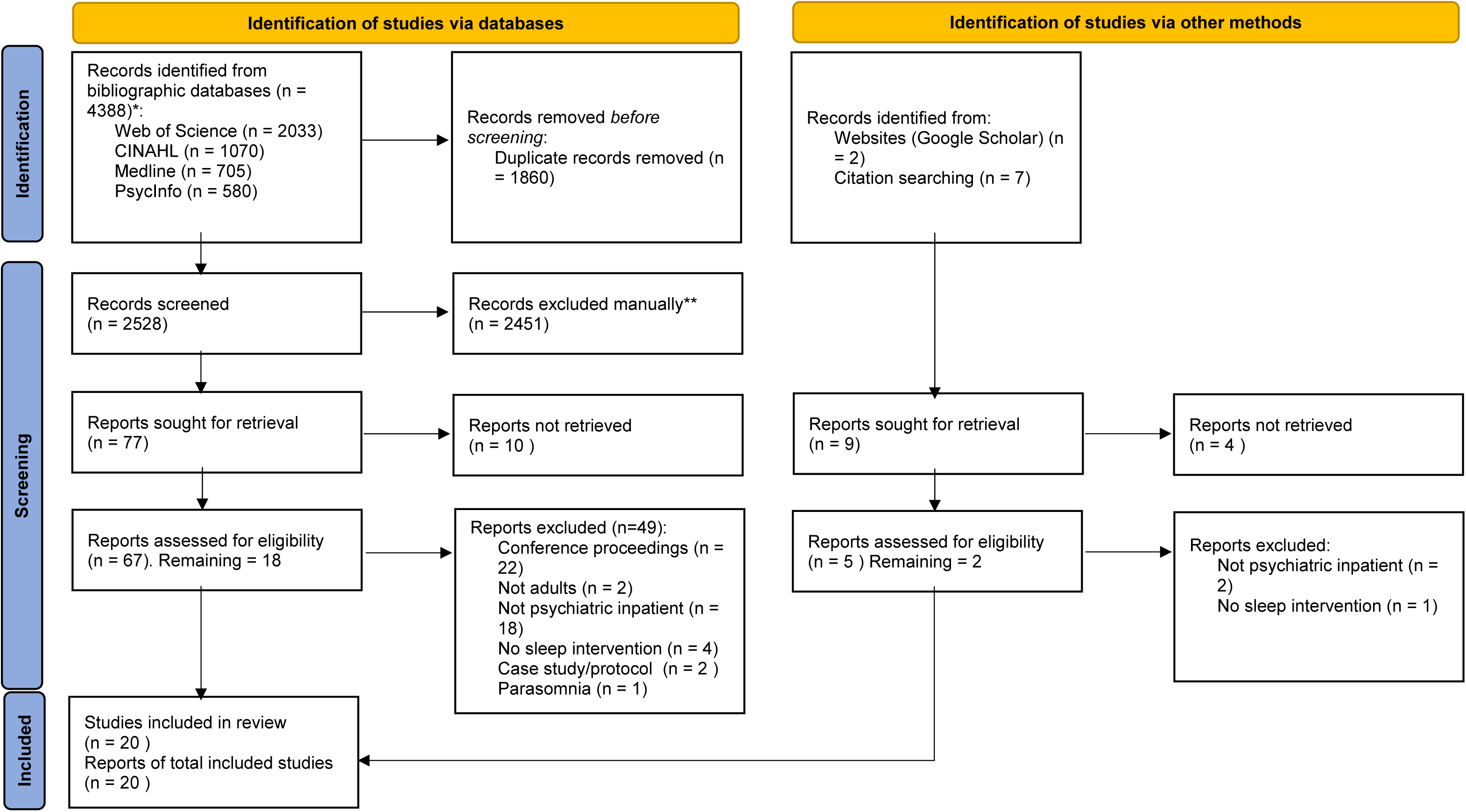
Adapted PRISMA flowchart of literature search and screening

### Study and participant characteristics

**Error! Not a valid bookmark self-reference.** summarises key features of the 20 included studies which were published between 1968 and 2020. All studies were conducted in high-income countries, spanning four continents: Europe (n = 13, 65%) (45–57), Asia (n = 3, 15%) (58–60) and North America (n = 3, 15%) (61–63) and Australia (n = 1, 5%) (64). One-third (n=7) of studies were RCTs (52, 55–58, 60, 61). Of the remaining quasi-experimental or non-randomised studies, six included a control group (46, 48, 49, 51, 65, 66). The mean MMAT score of quality was 85%. The risk of bias across the RCTs is presented in Supplement 3.

**Table 1:**
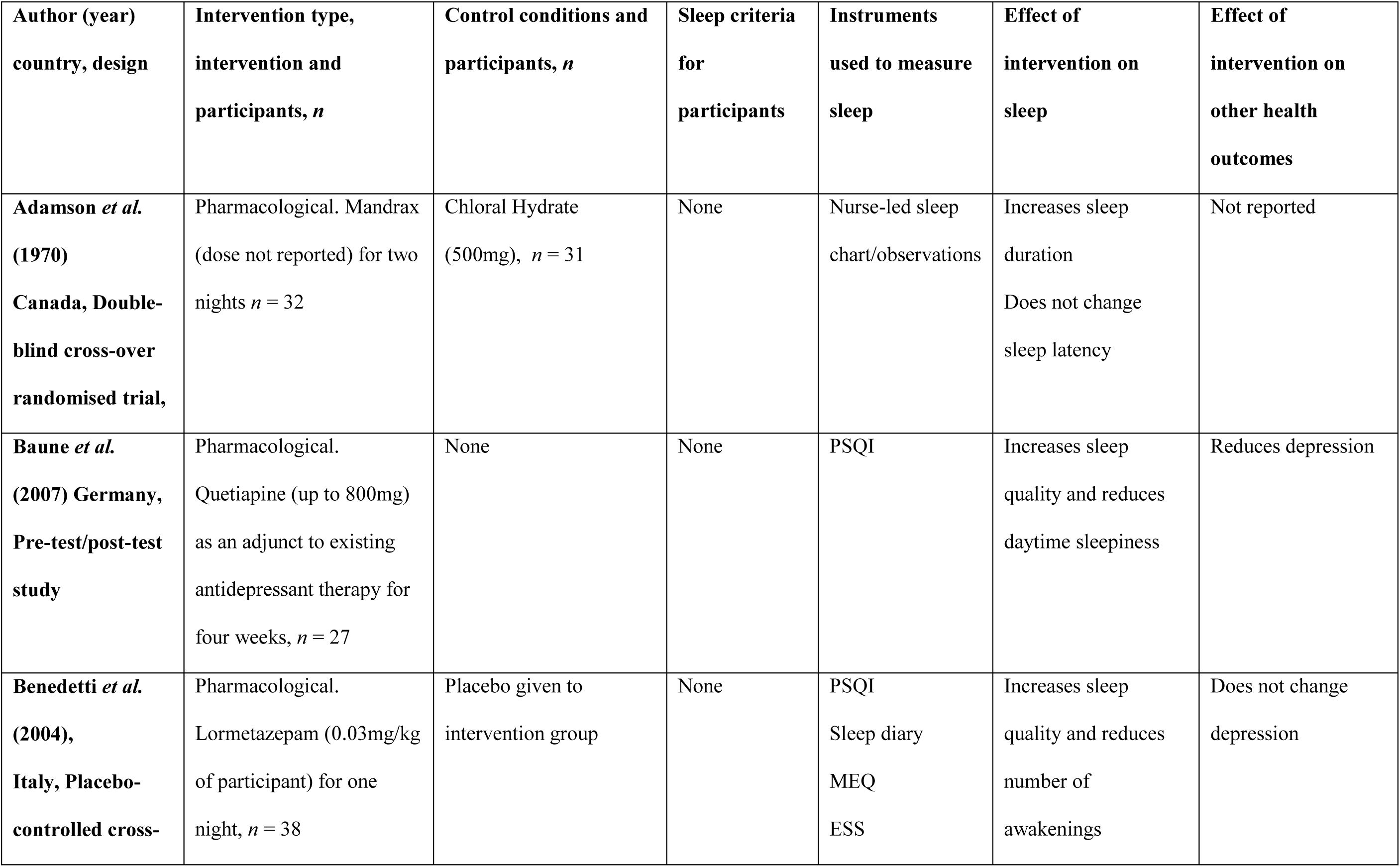
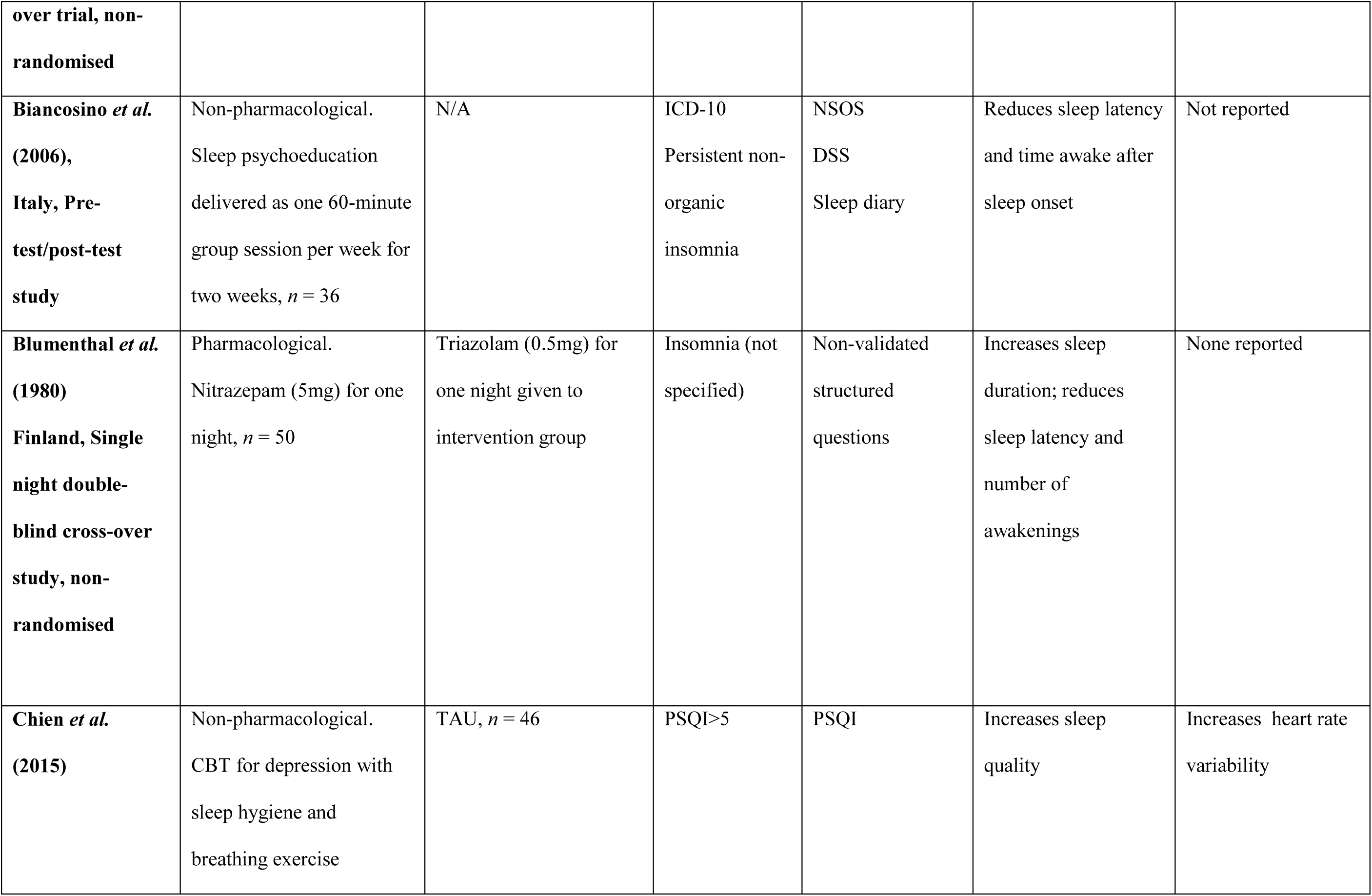
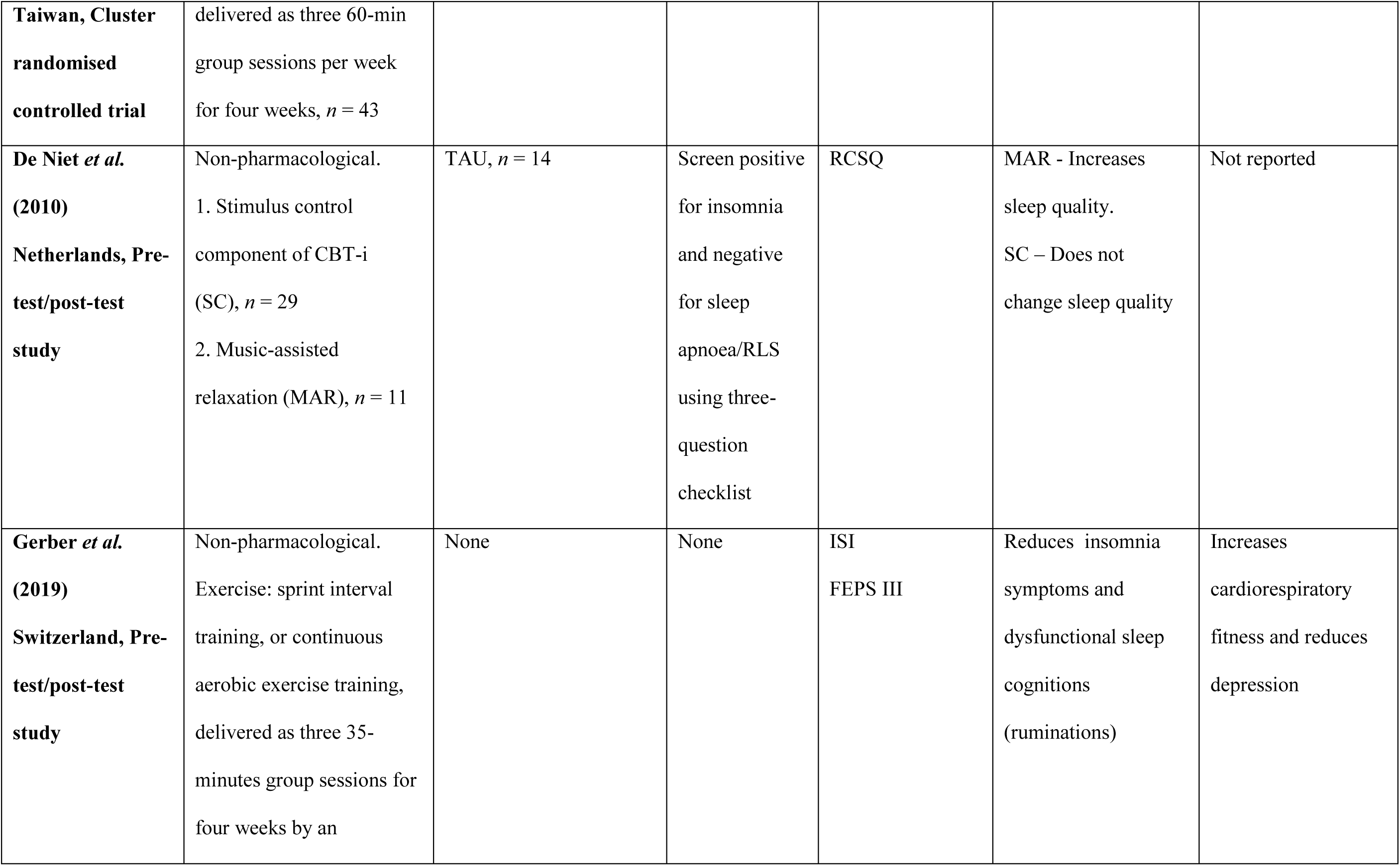
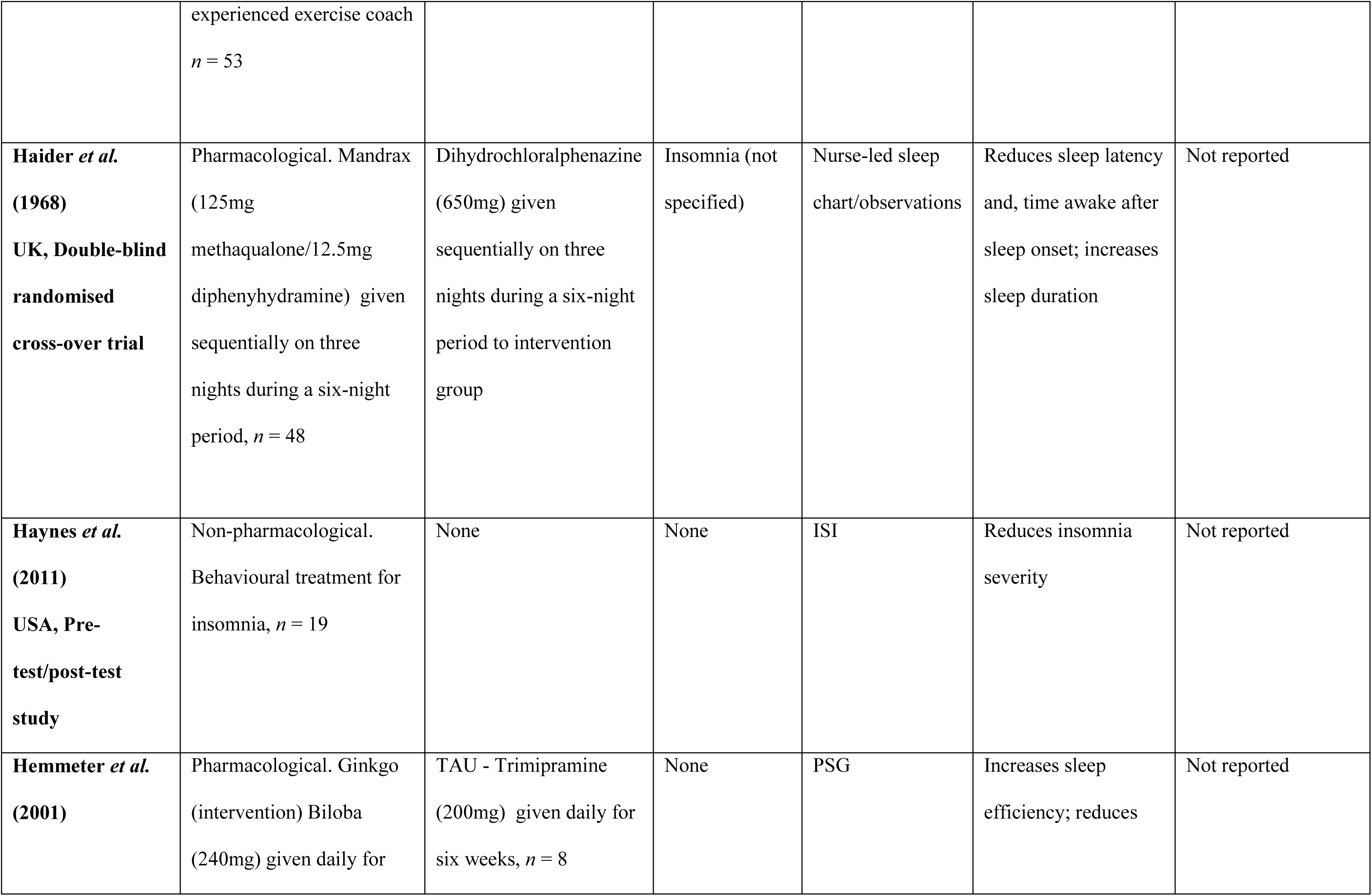
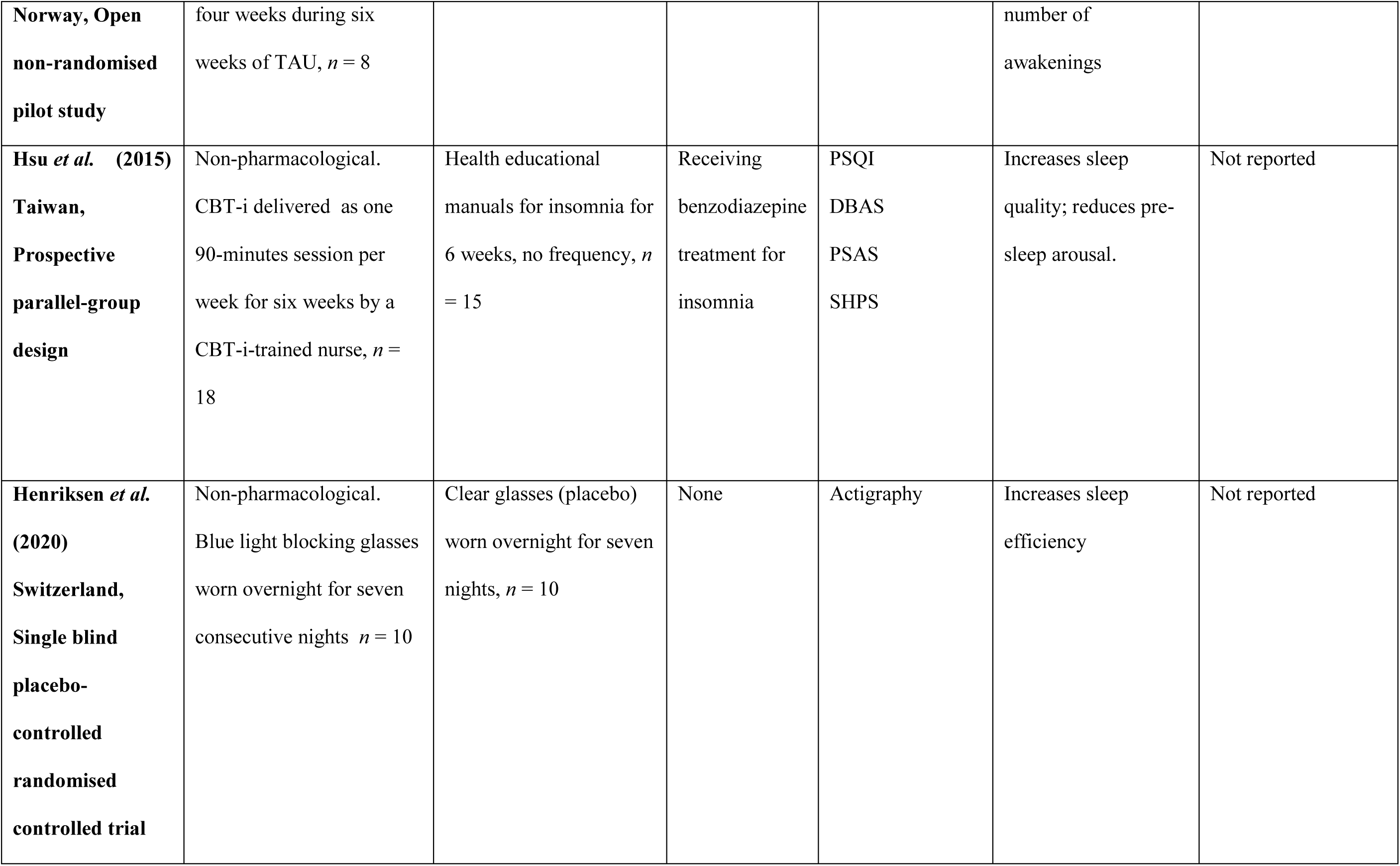
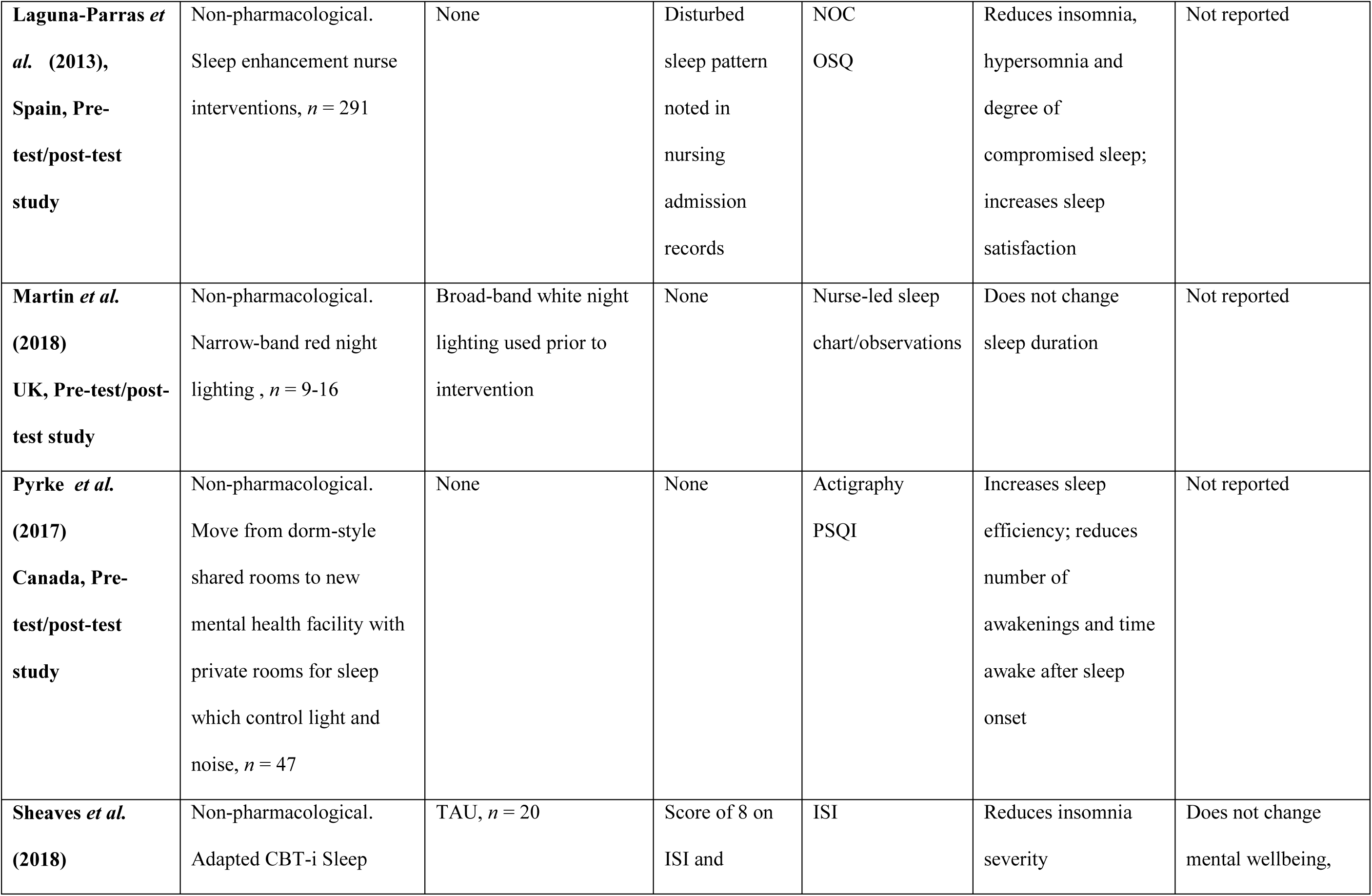
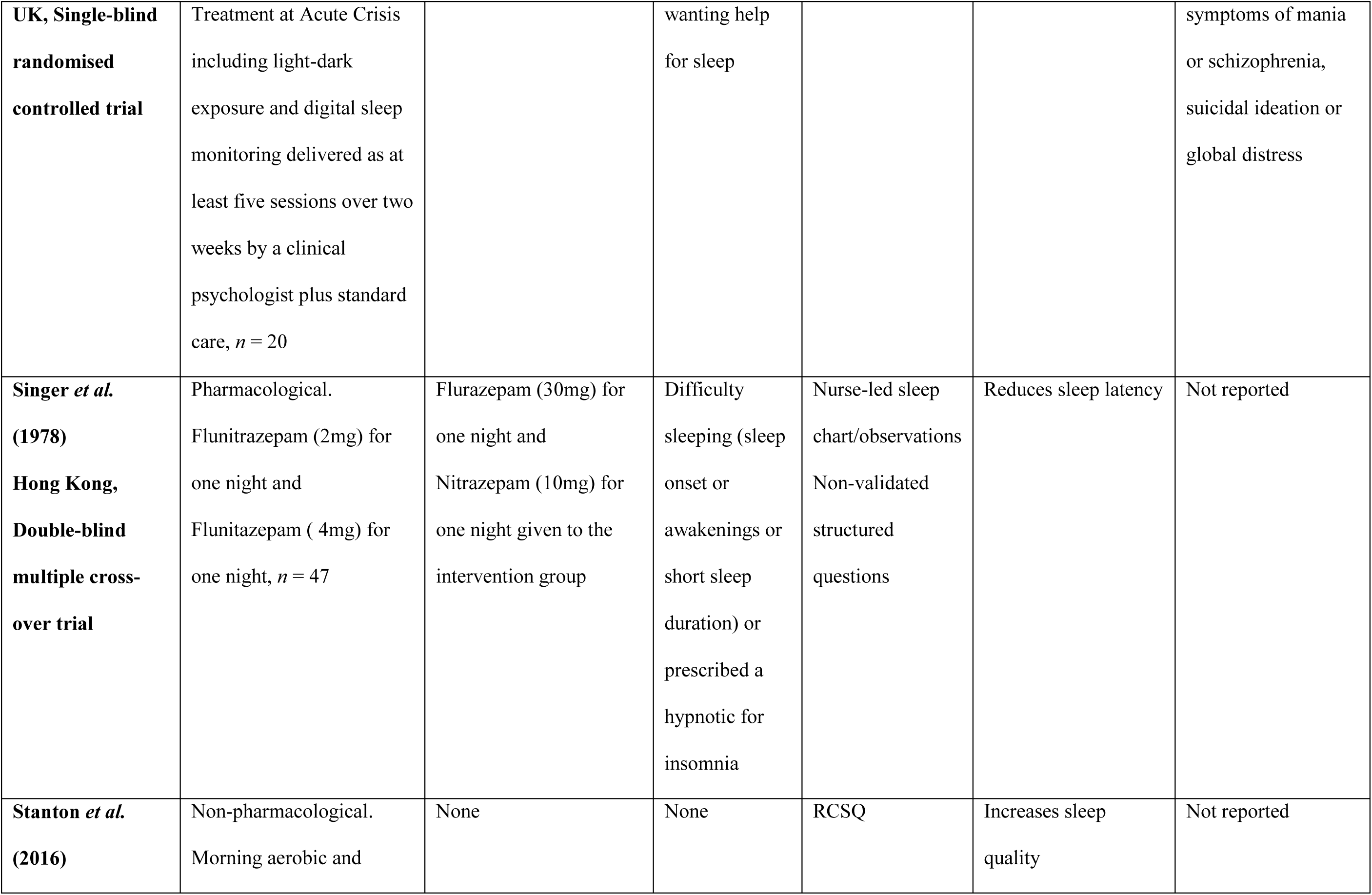
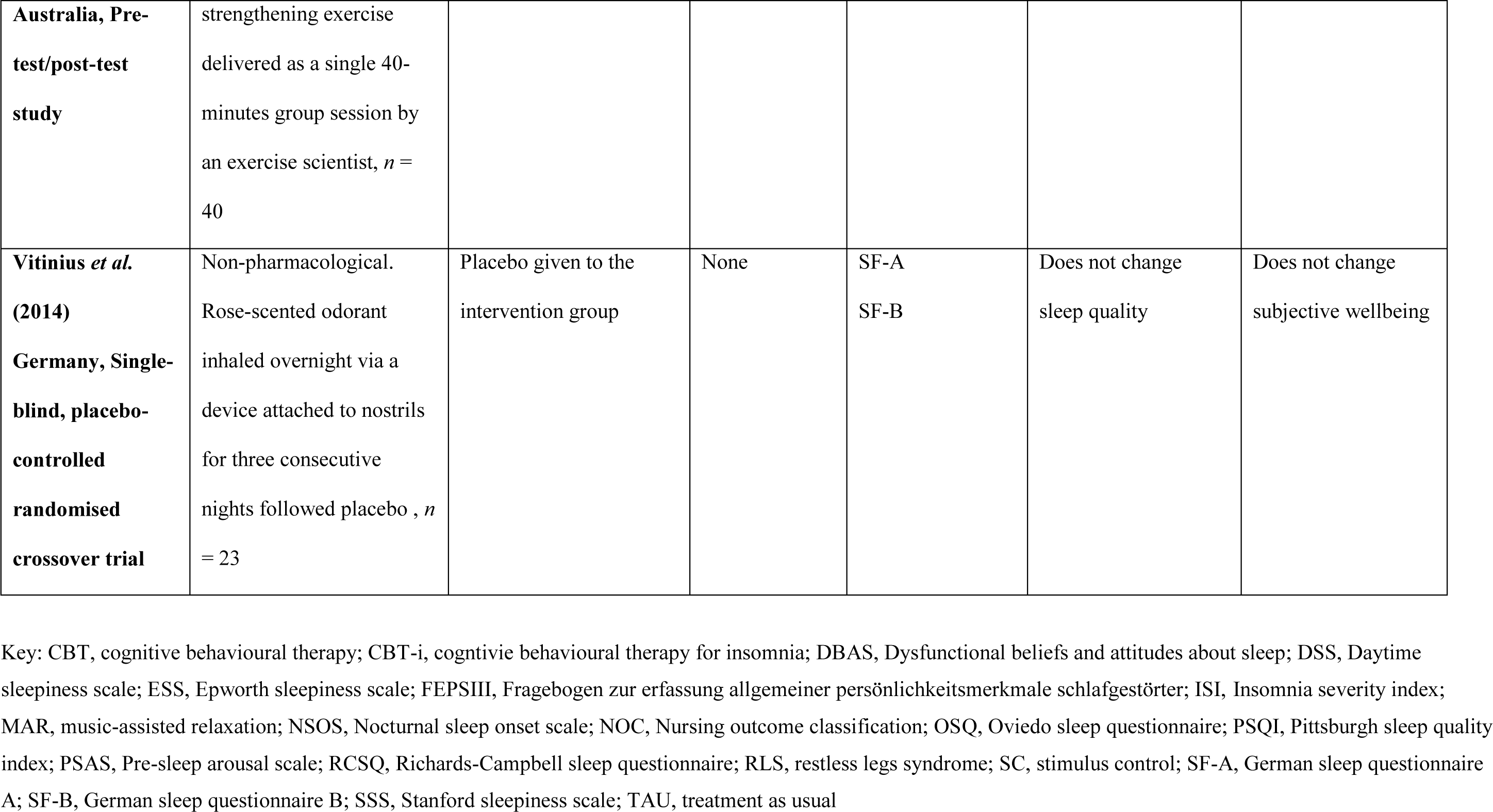
Characteristics and main findings of included studies

| **Author (year)**<br>**country, design** | **Intervention type,**<br>**intervention and**<br>**participants, *n*** | **Control conditions and**<br>**participants, *n*** | **Sleep criteria**<br>**for**<br>**participants** | **Instruments**<br>**used to measure**<br>**sleep** | **Effect of**<br>**intervention on**<br>**sleep** | **Effect of**<br>**intervention on**<br>**other health**<br>**outcomes** |
| --- | --- | --- | --- | --- | --- | --- |
| **Adamson *et al.***<br>**(1970)**<br>**Canada, Double-**<br>**blind cross-over**<br>**randomised trial,** | Pharmacological. Mandrax<br>(dose not reported) for two<br>nights *n* = 32 | Chloral Hydrate<br>(500mg), *n* = 31 | None | Nurse-led sleep<br>chart/observations | Increases sleep<br>duration<br>Does not change<br>sleep latency | Not reported |
| **Baune *et al.***<br>**(2007) Germany,**<br>**Pre-test/post-test**<br>**study** | Pharmacological.<br>Quetiapine (up to 800mg)<br>as an adjunct to existing<br>antidepressant therapy for<br>four weeks, *n* = 27 | None | None | PSQI | Increases sleep<br>quality and reduces<br>daytime sleepiness | Reduces depression |
| **Benedetti *et al.***<br>**(2004),**<br>**Italy, Placebo-**<br>**controlled cross-** | Pharmacological.<br>Lormetazepam (0.03mg/kg<br>of participant) for one<br>night, *n* = 38 | Placebo given to<br>intervention group | None | PSQI<br>Sleep diary<br>MEQ<br>ESS | Increases sleep<br>quality and reduces<br>number of<br>awakenings | Does not change<br>depression |
| **over trial, non-randomised** |  |  |  |  |  |  |
| **Biancosino *et al.* (2006), Italy, Pre-test/post-test study** | Non-pharmacological. Sleep psychoeducation delivered as one 60-minute group session per week for two weeks, *n* = 36 | N/A | ICD-10 Persistent non-organic insomnia | NSOS DSS Sleep diary | Reduces sleep latency and time awake after sleep onset | Not reported |
| **Blumenthal *et al.* (1980) Finland, Single night double-blind cross-over study, non-randomised** | Pharmacological. Nitrazepam (5mg) for one night, *n* = 50 | Triazolam (0.5mg) for one night given to intervention group | Insomnia (not specified) | Non-validated structured questions | Increases sleep duration; reduces sleep latency and number of awakenings | None reported |
| **Chien *et al.* (2015)** | Non-pharmacological. CBT for depression with sleep hygiene and breathing exercise | TAU, *n* = 46 | PSQI>5 | PSQI | Increases sleep quality | Increases heart rate variability |
| **Taiwan, Cluster randomised controlled trial** | delivered as three 60-min group sessions per week for four weeks, $n = 43$ | | | | | |
| **De Niet *et al.* (2010)**<br>**Netherlands, Pre-test/post-test study** | Non-pharmacological.<br>1. Stimulus control component of CBT-i (SC), $n = 29$<br>2. Music-assisted relaxation (MAR), $n = 11$ | TAU, $n = 14$ | Screen positive for insomnia and negative for sleep apnoea/RLS using three-question checklist | RCSQ | MAR - Increases sleep quality.<br>SC – Does not change sleep quality | Not reported |
| **Gerber *et al.* (2019)**<br>**Switzerland, Pre-test/post-test study** | Non-pharmacological.<br>Exercise: sprint interval training, or continuous aerobic exercise training, delivered as three 35-minutes group sessions for four weeks by an experienced exercise coach<br>*n* = 53 | None | None | ISI<br>FEPS III | Reduces insomnia symptoms and dysfunctional sleep cognitions (ruminations) | Increases cardiorespiratory fitness and reduces depression |
| **Haider *et al.***<br>**(1968)**<br>**UK, Double-blind**<br>**randomised**<br>**cross-over trial** | Pharmacological. Mandrax<br>(125mg<br>methaqualone/12.5mg<br>diphenhydramine) given<br>sequentially on three<br>nights during a six-night<br>period, *n* = 48 | Dihydrochloralphenazine<br>(650mg) given<br>sequentially on three<br>nights during a six-night<br>period to intervention<br>group | Insomnia (not<br>specified) | Nurse-led sleep<br>chart/observations | Reduces sleep latency<br>and, time awake after<br>sleep onset; increases<br>sleep duration | Not reported |
| **Haynes *et al.***<br>**(2011)**<br>**USA, Pre-**<br>**test/post-test**<br>**study** | Non-pharmacological.<br>Behavioural treatment for<br>insomnia, *n* = 19 | None | None | ISI | Reduces insomnia<br>severity | Not reported |
| **Hemmeter *et al.***<br>**(2001)** | Pharmacological. Ginkgo<br>(intervention) Biloba<br>(240mg) given daily for | TAU - Trimipramine<br>(200mg) given daily for<br>six weeks, *n* = 8 | None | PSG | Increases sleep<br>efficiency; reduces | Not reported |
| **Norway, Open non-randomised pilot study** | four weeks during six weeks of TAU, $n = 8$ | | | | number of awakenings | |
| **Hsu *et al.* (2015)**<br>**Taiwan,**<br>**Prospective parallel-group design** | Non-pharmacological. CBT-i delivered as one 90-minutes session per week for six weeks by a CBT-i-trained nurse, $n = 18$ | Health educational manuals for insomnia for 6 weeks, no frequency, $n = 15$ | Receiving benzodiazepine treatment for insomnia | PSQI<br>DBAS<br>PSAS<br>SHPS | Increases sleep quality; reduces pre-sleep arousal. | Not reported |
| **Henriksen *et al.* (2020)**<br>**Switzerland,**<br>**Single blind placebo-controlled randomised controlled trial** | Non-pharmacological. Blue light blocking glasses worn overnight for seven consecutive nights $n = 10$ | Clear glasses (placebo) worn overnight for seven nights, $n = 10$ | None | Actigraphy | Increases sleep efficiency | Not reported |
| **Laguna-Parras *et al.* (2013), Spain, Pre-test/post-test study** | Non-pharmacological.<br>Sleep enhancement nurse interventions, *n* = 291 | None | Disturbed sleep pattern noted in nursing admission records | NOC<br>OSQ | Reduces insomnia, hypersomnia and degree of compromised sleep; increases sleep satisfaction | Not reported |
| **Martin *et al.* (2018) UK, Pre-test/post-test study** | Non-pharmacological.<br>Narrow-band red night lighting , *n* = 9-16 | Broad-band white night lighting used prior to intervention | None | Nurse-led sleep chart/observations | Does not change sleep duration | Not reported |
| **Pyrke *et al.* (2017) Canada, Pre-test/post-test study** | Non-pharmacological.<br>Move from dorm-style shared rooms to new mental health facility with private rooms for sleep which control light and noise, *n* = 47 | None | None | Actigraphy<br>PSQI | Increases sleep efficiency; reduces number of awakenings and time awake after sleep onset | Not reported |
| **Sheaves *et al.* (2018)** | Non-pharmacological.<br>Adapted CBT-i Sleep | TAU, *n* = 20 | Score of 8 on ISI and | ISI | Reduces insomnia severity | Does not change mental wellbeing, |
| **UK, Single-blind randomised controlled trial** | Treatment at Acute Crisis including light-dark exposure and digital sleep monitoring delivered as at least five sessions over two weeks by a clinical psychologist plus standard care, *n* = 20 |  | wanting help for sleep |  |  | symptoms of mania or schizophrenia, suicidal ideation or global distress |
| **Singer *et al.* (1978)**<br>**Hong Kong, Double-blind multiple cross-over trial** | Pharmacological.<br>Flunitrazepam (2mg) for one night and<br>Flunitazepam ( 4mg) for one night, *n* = 47 | Flurazepam (30mg) for one night and<br>Nitrazepam (10mg) for one night given to the intervention group | Difficulty sleeping (sleep onset or awakenings or short sleep duration) or prescribed a hypnotic for insomnia | Nurse-led sleep chart/observations<br>Non-validated structured questions | Reduces sleep latency | Not reported |
| **Stanton *et al.* (2016)** | Non-pharmacological.<br>Morning aerobic and | None | None | RCSQ | Increases sleep quality | Not reported |
| **Australia, Pre-test/post-test study** | strengthening exercise delivered as a single 40-minutes group session by an exercise scientist, $n = 40$ | | | | | |
| **Vitinius *et al.* (2014)**<br>**Germany, Single-blind, placebo-controlled randomised crossover trial** | Non-pharmacological. Rose-scented odorant inhaled overnight via a device attached to nostrils for three consecutive nights followed placebo, $n = 23$ | Placebo given to the intervention group | None | SF-A<br>SF-B | Does not change sleep quality | Does not change subjective wellbeing |

Key: CBT, cognitive behavioural therapy; CBT-i, cognitive behavioural therapy for insomnia; DBAS, Dysfunctional beliefs and attitudes about sleep; DSS, Daytime sleepiness scale; ESS, Epworth sleepiness scale; FEPSIII, Fragebogen zur erfassung allgemeiner persönlichkeitsmerkmale schlafgestörter; ISI, Insomnia severity index; MAR, music-assisted relaxation; NSOS, Nocturnal sleep onset scale; NOC, Nursing outcome classification; OSQ, Oviedo sleep questionnaire; PSQI, Pittsburgh sleep quality index; PSAS, Pre-sleep arousal scale; RCSQ, Richards-Campbell sleep questionnaire; RLS, restless legs syndrome; SC, stimulus control; SF-A, German sleep questionnaire A; SF-B, German sleep questionnaire B; SSS, Stanford sleepiness scale; TAU, treatment as usual

There has been a marked increase in the number of studies of sleep interventions in adult psychiatric inpatient settings published since 2011. Additional trends observed in these studies are highlighted in Supplement 4. The majority of included studies (n=17, 85%) were conducted in non-specialist inpatient settings with the remainder of studies conducted on wards for older adults (54), military personnel (48) or military veterans (62). Across all included studies, there were 1034 adults, with one study not clearly reporting the sample size (54). All but one study (53) (n=291 participants) reported fewer than 100 participants.

Among the RCTs, the size of the intervention group ranged from 10 (52) to 48 (57). Among the 15 studies that reported gender, 54% (n=452) of participants were male. The age of participants across the studies ranged from 18 years (48) to over 71 years.(54, 57)

Nine (45%) studies involved participants with mixed psychiatric diagnoses (47, 49, 53, 55, 57, 58, 61–63) and seven (35%) studies recruited participants who had a depressive disorder (45, 46, 50, 51, 56, 60, 65). The remaining studies included participants with a common diagnosis of mania (n=1) (52), mixed mood and anxiety disorder(n=1) (64) and dementia and cognitive impairment (n=1). One study did not report participant diagnosis (48).

### Measurement of sleep outcomes and barriers to measurement

There were 20 distinct instruments used to measure effects on sleep (Supplement 5). Three (15%) studies used objective measures of polysomnography (PSG) (51) and actigraphy (52, 63). Of the 15 validated subjective instruments, the two most frequently used were the Pittsburgh Sleep Quality Index (PSQI) (45, 46, 60, 63, 65) and the Insomnia Severity Index (ISI) (n=3) (50, 55, 62). Twelve (65%) studies used only one instrument (45, 48, 49, 51, 52, 54, 55, 57, 60–62, 64), whilst 40% (n=8) used at least two instruments (46, 47, 50, 52, 53, 56, 58, 63, 65), of which two combined an objective sleep measure (actigraphy) with a validated subjective sleep measure (52, 63). There has been a small increase in studies using objective sleep measures since 1968 and a marked reduction in studies using non-validated subjective measures such as nurse-led sleep charts and patient-reported sleep diaries (Supplement 4).

Two (10%) studies reported barriers to measuring sleep in psychiatric inpatient settings (52, 62) . Occasional invalid or failed readings was a reported barrier to using actigraphy (52). The ISI could not be completed accurately when a participant with schizophrenia was described as disoriented and not lucid (62).

### Sleep interventions used

Pharmacological interventions were used in seven (35%) studies (45, 46, 48, 51, 57, 58, 61). Of these, five studies examined the effects of benzodiazepine (46, 48, 58) or non-benzodiazepine hypnotics (57, 61), whilst two studies tested the impact of antidepressants on sleep (45, 51). The remaining studies (n=13, 65%) used non-pharmacological interventions. Of these studies, nine studies used interventions based on CBT-i. There was a high level of heterogeneity among these studies with only one study(65) using standard non-adapted CBT-i with all the core elements of CBT-i: (i) sleep restriction, (ii) psychoeducation/ sleep hygiene, (iii) stimulus control, (iv) relaxation and (v) cognitive therapy. The remaining CBT-i-based studies used additional components (55) or at least one, but not all, of the five core elements of CBT-i (47, 49, 50, 53, 60, 62, 64). Specifically, two studies tested interventions of physical activity (50, 64) which is advice offered in the sleep hygiene/psychoeducation component of CBT-i. Environmental interventions were used in four studies which tested the effects of room occupancy (63), light (52, 54) and odour (56) on sleep.

### Effects of pharmacological interventions

#### Studies with a control group

All but one of the studies of pharmacological interventions included a control group. Among these, benzodiazepine and non-benzodiazepine hypnotics increased sleep duration (48, 57, 61) and reduced sleep latency (48, 57, 58). Both types of hypnotic reduced the time awake after sleep onset (57), with benzodiazepines specifically reducing the number of awakenings (46, 48). Hypnotic medication increased sleep quality but did not change depression scores (46). The use of antidepressant Gingko Biloba alongside existing antidepressant therapy was shown via PSG to increase sleep efficiency by reducing the number of awakenings; the effect on depression was not reported (51).

#### Studies without a control group

In the pharmacological study that did not include a control group, Quetiapine (an antidepressant and antipsychotic) used alongside existing antidepressant therapy not only increased sleep quality and reduced daytime sleepiness, but also reduced depression (45). There were no reported effects of pharmacological interventions on physical health in the included studies.

### Effects of non-pharmacological interventions (cognitive behavioural – based)

#### Studies with a control group

Of the nine studies that involved interventions based on cognitive behaviour therapy, four (44.4%) included control groups (49, 55, 60, 65). Standard CBT-i increased sleep quality and reduced levels of pre-sleep arousal (65). With the addition of patient-worn digital devices to monitor sleep and increase motivation, light-dark exposure to enhance circadian rhythm and strategies to reduce the impact of observations by staff at night, CBT-i reduced insomnia symptoms. Despite the positive effect on sleep, this enhanced CBT-i did not change mental wellbeing, suicidality, manic symptoms or symptoms of schizophrenia (55). Whilst CBT-i was significantly more effective at improving sleep quality than sleep hygiene/ psychoeducation (65), there was little evidence that sleep psychoeducation alone improved sleep (49). However, sleep hygiene/psychoeducation administered with breathing exercises and CBT for depression not only increased sleep quality but had the added physical health benefit of increasing heart rate variability (60). (Lower heart rate variability predicts higher all-cause mortality (67).) Subjective sleep quality increased when sleep hygiene/psychoeducation was supplemented with music-assisted relaxation but not when given alone or with stimulus control (49).

#### Studies without a control group

Five non-pharmacological studies based on cognitive behavioural therapy had a single-arm design with no control (47, 50, 53, 62, 64). Of these, two focused exclusively on exercise (50, 64). In contrast to several pharmacological interventions (46, 48, 51), a non-pharmacological intervention with three components of CBT-i (sleep hygiene/psychoeducation, stimulus control and sleep restriction) did not reduce the number of awakenings (47). However, it was associated with reduced sleep latency, reduced time awake after sleep onset, and reduced use of as required insomnia medication (47). An intervention comprising four of the six components of standard CBT-i (sleep hygiene, stimulus control, relaxation and cognitive therapy) reduced insomnia (62). Moreover, a similar intervention offering five CBT-i components not only reduced insomnia, but also reduced hypersomnia and increased sleep satisfaction (53). Sleep interventions involving only guided physical exercise increased sleep quality (64), reduced insomnia and were associated with fewer dysfunctional sleep cognitions (50). Additional mental and physical health benefits of exercise (interval training or aerobic exercise) reduced depression and increased cardiorespiratory fitness, respectively (50).

### Effects of non-pharmacological interventions (environmental)

#### Studies with a control group

Three (75%) environmental intervention studies included a comparison group (52, 54, 56). An RCT compared clear glasses with “blue-blocking” glasses which block the low wavelength blue light that suppresses melatonin (52). Using actigraphy, the study found that wearing blue-blocking glasses between 18:00 and 08:00 increased sleep efficiency (52). In contrast, another chronotherapy intervention (switching night lights in the hospital from white light - which includes blue and red light - to high wavelength red light) did not change sleep duration as measured through nursing observations (54). Sleeping in a room with a rose odour did not change sleep quality and had no effect on wellbeing (56).

#### Studies without a control group

An intervention-only study found that a hospital move from shared to single bedroom accommodation improved actigraphy-measured sleep efficiency by reducing the time awake after sleep onset and reducing the number of awakenings (63). However, the change of accommodation did not reduce total sleep time or perceived sleep quality (63).

## Discussion

Over the past 50 years, there has been an increase in sleep intervention studies undertaken in psychiatric inpatient settings. Reviews have been published that combine studies from psychiatric inpatient settings with those from non-psychiatric inpatient settings (33), prisons (35) and psychiatric community settings (68). To our knowledge, this is the first scoping review of sleep intervention studies of adults with mental disorders admitted only to psychiatric wards. The review findings showed that most studies focused on non-pharmacological than pharmacological interventions in the psychiatric inpatient setting. Furthermore, non-pharmacological sleep interventions largely improved sleep and had the potential for improving other health outcomes. Most studies of pharmacological interventions were RCTs whereas many studies of non-pharmacological interventions did not include a comparison group. The use of objective sleep measures was limited and subjective assessment tools varied considerably making the validity of findings uncertain. Studies rarely reported barriers to measuring sleep in the psychiatric inpatient setting.

### Measurement of sleep

Instruments used to measure sleep varied and were mostly validated subjective questionnaires. The most common was the PSQI (69). Over time, fewer studies have relied on non-validated subjective measures like sleep diaries and nursing observations. Our review identified a lack of studies using objective sleep measurements including polysomnography (51) and actigraphy (52, 63), in line with another review on sleep in prison (70).

Polysomnography is considered the gold standard in sleep medicine (71, 72). However, compared to subjective measurements, these technologies are costly and often difficult to access and implement in clinical settings (69). Objective measures do not require patients to have the level of cognitive functioning that is necessary for the use of subjective sleep questionnaires (49, 62). However, objective sleep measurements involving the use of batteries or wires may be less suitable for use with patients at high risk of self-harm and suicide. Whilst patient-reported subjective measurements encourage positive patient involvement, they can underestimate sleep duration even when objectively sleep duration is normal (73). This means it is possible that even when sleep is objectively improved with mental health benefits, patients may subjectively perceive that they are not sleeping better. In selecting a sleep measurement, consideration should be given to the degree to which the measurement is validated to measure the specific process of sleep or circadian rhythm that the researcher or clinician intends to measure. For example, actigraphy provides a reliable measurement of daytime activity and is highly sensitive to sleep, but overestimates total sleep time and is less effective in measuring circadian rhythm (71, 72). Similarly, the degree to which a patient feels sufficiently rested on waking to get up and start the day is better assessed using subjective rather than objective measurements. Whilst the increased use of objective measures is recommended in psychiatric inpatient setting, they are not always appropriate used alone and should therefore be complemented, or in some cases replaced, by subjective measures which ideally should be validated. Ultimately, the choice of sleep measurement will be guided by the sleep process intended to be measured, individual patient factors and financial resources.

### Interventions

Our review identified many studies reporting effective pharmacological and non-pharmacological sleep interventions in psychiatric inpatient settings among patients with a diversity of mental disorders. However, a few studies did not find any sleep benefits. No studies reported adverse effects of an intervention on sleep. In comparison, a larger meta-analysis of RCTs of sleep interventions in prisons and psychiatric hospitals also found that the majority of studies reported positive effects on sleep, whereas 2% found adverse effects on sleep (35).

Findings were consistent with other studies showing the wide used of non-pharmacological interventions, particularly those based on CBT-i in psychiatric inpatient research (33, 35). CBT-i is the recommended first-line treatment for adults with chronic insomnia (74).

However, some patients in psychiatric inpatients settings will not be able to access or benefit from this intervention. For example, CBT-i may not be readily available in some psychiatric inpatient settings due to financial costs and lack of training (75). CBT-i also requires a high level of patient engagement and without this, the benefits are unlikely to be obtained.

Where financial resources are limited, a digital version of CBT-i (dCBT-i) could be used (76). DCBT-i has been used with adults experiencing mood and anxiety disorders (77) and is more cost-effective than individual and group face-to-face CBT-i as well as pharmacological sleep interventions (78). However, studies are needed to measure the effectiveness of and identify the barriers to using dCBT-i in the psychiatric hospital setting. Alternatively, in the absence of standard CBT-i, patients may still obtain some benefit from receiving one or more of the six components of CBT-i, such as psychoeducation with music-assisted relaxation (49).

CBT-i may be less suitable for some patients on psychiatric wards who lack motivation and concentration due to the nature and severity of their mental disorder (71). In such cases, there are effective pharmacological interventions for patients with capacity and a willingness to accept medication. Few studies reported effectiveness of environmental interventions such as blocking out blue light (52) on sleep among adults in the psychiatric inpatient setting. Therefore, more randomised controlled trials are needed before implementation is possible. Environmental interventions may offer an alternative to medication for patients unable to benefit from CBT-i.

Some studies showed sleep interventions have potential to improve sleep whilst also improving physical fitness and reducing mental ill-health. This is in line with evidence from healthy and other clinical populations on the direct associations between improved sleep and physical health (12), more positive affect (79), improved cognition (22), reduced suicidality (23) and reduced aggression (25). However, more controlled studies are needed with large sample sizes in psychiatric inpatient settings to examine the impact of sleep interventions on other health outcomes.

## Limitations

To complement existing reviews, our search strategy included a larger number of databases, excluded studies that were not conducted in psychiatric inpatient settings (35), and did not exclude pharmacological interventions (33). However, we did not search grey literature which may have identified additional studies. In restricting the review to studies published in English, we excluded sleep interventions described in other languages. The risk of selection bias could have been reduced by using two reviewers instead of a single reviewer to screen titles and abstracts. A key shortcoming is the relatively low proportion of non-pharmacological studies that were designed without a comparison group. This limited the opportunity to draw conclusions about the effectiveness of many non-pharmacological sleep interventions.

There was a disproportionately high number of studies from European countries with no representation from low and middle income countries (LMICs), despite a growing number of sleep health publications outside of high income countries (80, 81). Whilst the prevalence of sleep disturbances does not appear to vary globally (81), some LMICs have unique cultural understandings of sleep (80), which could affect the acceptability of some sleep interventions that are used effectively in high income countries. This review did not aim to identify interventions that report cost-effectiveness within a psychiatric inpatient studies, but this information would be particularly useful for LMICs.

## Conclusions

This review has identified a growing body of evidence for the use of non-pharmacological and, to a lesser degree, pharmacological, interventions to improve sleep of adults in psychiatric inpatient settings. Objective sleep measures were limited and rarely used in these settings. Validated subjective measures can complement objective measures used in inpatient psychiatric sleep research. The review highlights gaps in the evidence for environmental sleep interventions (e.g. chronotherapy), from research conducted low and middle income countries and from studies that measure additional health outcomes. There is a need for more randomised controlled trials into transdiagnostic sleep interventions that can be used in adults in psychiatric inpatient settings.

### Practice points

1. Objective sleep measures (eg, polysomnography and actigraphy) should be used where possible in future studies and should be complemented by validated subjective measures (eg, PSQI and ISI).
2. Clinicians can choose from a range of validated objective and subjective sleep measures instead of relying on non-validated subjective measures to assess the sleep in adults admitted to the psychiatric setting.

### Research agenda

To advance research into sleep interventions for adults with mental disorders in inpatient psychiatric settings:

1. There is a need for further research into the effect of environmental interventions on sleep.
2. More evidence of the effectiveness of transdiagnostic sleep interventions is needed.
3. Study designs should include a comparison group.
4. There is a need for sleep intervention research conducted in psychiatric inpatients settings in low and middle income countries.
5. Longitudinal studies are needed to understand any distal effects of sleep interventions on mental and physical health.
6. Greater homogeneity of reported sleep outcomes is desired between intervention Studies

## Supporting information

Supplement 1 - search strategy

Supplement 2 - GRIPP2

Supplement 3 - risk of bias

Supplement 4 - trends since 1968

Supplement 5 - instruments used to measure sleep

## Data Availability

All data produced in the present study are available upon reasonable request to the authors

## Abbreviations

CBT-i: Cognitive behavioural therapy for insomnia
dCBT-i: Digital cognitive behavioural therapy for insomnia
GRIPP2: Guidance for reporting involvement of patients and the public version 2
ISI: Insomnia severity index
JBI: Joanna Briggs institute
LMICs: Low and middle income countries
MMAT: Mixed methods appraisal tool
PSQI: Pittsburgh sleep quality index
PRISMA: Preferred reporting items for systematic reviews and meta-analyses
PRISMA-ScR: Preferred reporting items for systematic reviews and meta-analyses extension for scoping reviews
RCT: Randomised controlled trial

## Acknowledgements

The authors acknowledge contributions made at the start of the study by Mr. B, a Forensic Sleep Research Group member with lived experience of the scoping review topic. We thank the Information Service Manager at the Centre for Reviews and Dissemination, University of York, UK for undertaking searches on databases. We also acknowledge the role of a Specialty Trainee in Forensic Psychiatry, Tees, Esk and Wear Valleys NHS Foundation Trust, UK who contributed to the scoping review protocol. An.Ab received funding from the NIHR Applied Research Collaboration North East & North Cumbria, UK, to support writing the final two drafts of the manuscript.

