## Supplement 1 - search strategy for "Sleep interventions for adults admitted to psychiatric inpatient settings: a systematic scoping review"

| Search – OVID Medline 1946 to September 18, 2019 on 20 September 2019 | Query | Records retrieved |
| --- | --- | --- |
| #1 | sleep.m_titl. OR "sleep-less*".m_titl. OR "sleep problem*".m_titl. OR "sleep disturbance*".m_titl. OR poor sleep.m_titl. OR "hypnotic*".m_titl. OR z-drug.m_titl. OR zopiclone.m_titl. OR zolpidem.m_titl. OR zaleplon.m_titl. OR melatonin.m_titl. OR insomnia.m_titl. OR insomniac.m_titl. OR *somn*".m_titl. OR "actigraph*".m_titl. OR "actimetr*".m_titl. OR "accelerometer*".m_titl. OR exp Sleep Disorders, Intrinsic/ or exp Sleep, Slow-Wave/ or exp Sleep Hygiene/ or exp Sleep Stages/ or exp Sleep Deprivation/ or exp Sleep/ or exp Sleep Wake Disorders/ or exp Sleep, REM/ or exp Sleep Disorders, Circadian Rhythm/ OR exp "Hypnotics and Sedatives"/ OR exp Receptors, Melatonin/ or exp Melatonin/ or exp Receptor, Melatonin, MT2/ or exp Receptor, Melatonin, MT1/ OR exp "Sleep Initiation and Maintenance Disorders"/ OR Polysomnography/ OR Accelerometry/ \| "sleep-less*".m_titl. | 288968 |
| #2 | (mental or psychiatr* or psychos* or psychot* or schizo* or depressi* or depressed or personality disorder or personality disorders or bipolar or mood disorder or mood disorders or affective disorder or affective disorders).m_titl. OR exp Mental Health/ OR exp Schizophrenia/ OR exp Psychotic Disorders/ OR exp Depression, Postpartum/ or exp Depression/ OR Bipolar Disorder/ OR Personality Disorders/ | 639034 |
| #3 | "hospital*".m_titl. OR \| "unit*".m_titl. OR inpatient.m_titl. OR in-patient.m_titl. OR ward*.m_titl. OR PICU.m_titl. OR \| seclusion.m_titl. OR rehab*.m_titl. OR forensic*.m_titl. OR secure.m_titl. OR exp Hospitals, Psychiatric/ OR exp Hospitalization/ OR exp Inpatients/ OR exp Psychiatric Department, Hospital/ | 725718 |
| #4 | 1 AND 2 AND 3 AND 4 | 718 |
| No language and date limits were applied | |  |
