## Supplement 2 - GRIPP2 for "Sleep interventions for adults admitted to psychiatric inpatient settings: a systematic scoping review"

**Supplement 2: GRIPP2 – short form**

| **Section and topic** | **Item** | **Reported on page no.** |
| --- | --- | --- |
| **1: Aim** | The review focuses on adult patients with mental disorders admitted to the psychiatric inpatient setting. The aim of PPI in the scoping review was to ensure that the review topic:  (i) was informed by adults with lived experience of sleeping in the psychiatric inpatient setting and,  (ii) would achieve results to inform research that could have an impact on adults admitted to this setting. | 1, 7 |
| **2: Methods** | The Forensic Sleep Research Group was established following a meeting with presentations by sleep clinicians, sleep academics and patients admitted to psychiatric wards who shared their experiences of sleep in this setting. Further engagement work with a larger number of adults admitted to psychiatric wards confirmed that sleep was an important topic, deserving of further study.  One of the members of the Forensic Research Group is an adult with lived experience of sleeping in a psychiatric hospital. All members of the Forensic Sleep Research Group agreed the idea for the scoping review. Progress of the scoping review was discussed in these group meetings. The patient member received INVOLVE payments for their contributions. | 1 |
| **3: Study results** | The group member with lived experience was able to make contributions in the early stages of the scoping review (review topic, design of method for screening articles by title and abstract) and described feeling involved in the project. However, for several reasons, this group member was not able to attend subsequent meetings. The patient member of the Forensic Research Group was invited to contribute to the discussion and recommendations. | 1 |
| **4: Discussion and conclusions** | Whilst patients initially heavily drove the choice of study topic, they had relatively little involvement during the delivery and analysis stages of the review. | 1 |
| **5: Reflections/critical perspective** | During the pandemic, virtual meetings, which were less easy for the patient member of the group to attend, replaced face-to-face meetings. Holding hybrid meetings would have overcome this challenge and enable the patient member to remain more involved during the middle and later stages of the review. Furthermore, the Forensic Sleep Research Group might have benefited from having three patient members to ensure that there would be patient representation at every meeting, | N/A |
