## Supplement 3 - risk of bias for "Sleep interventions for adults admitted to psychiatric inpatient settings: a systematic scoping review"

**Supplement 3: Risk of bias summary for RCTs**

|  | Low risk of bias | Unclear risk of bias | High risk of bias |
| --- | --- | --- | --- |
| Other bias | 2 | 5 | 0 |
| Attrition bias | 3 | 3 | 1 |
| Detection bias | 1 | 6 | 0 |
| Performance bias | 2 | 5 | 0 |
| Reporting bias | 3 | 3 | 1 |
| Selection bias: allocation concealment | 2 | 5 | 0 |
| Selection bias: random sequence allocation | 2 | 5 | 0 |
