## Supplement 4 - trends since 1968 for "Sleep interventions for adults admitted to psychiatric inpatient settings: a systematic scoping review"

**Supplement 4: Trends in study characteristics between 1968 and 2020**

| **Study characteristics** | **Year of publication** | | |
| --- | --- | --- | --- |
|  | **1968 - 2000**  **N=4 studies** | **2001 - 2010**  **N=5 studies** | **2011 - 2020**  **N=11 studies** |
| European country of origin | 2 (50%) | 5 (100%) | 7 (64%) |
| RCT design | 3 (75%) | 0 (0%) | 4 (36%) |
| High quality based on MMAT score | 2 (50%) | 4 (80%) | 5 (45%) |
| Objective sleep measurement used | 0 (0%) | 1 (20%) | 2 (18%) |
| Any subjective non-validated measurement used | 4 (100%) | 2 (40%) | 1 (9%) |
| Only subjective non-validated measurement used | 4 (100%) | 0 (0%) | 1 (9%) |
| Pharmacological intervention tested | 4 (100%) | 3 (60%) | 0 (0%) |
| Non-pharmacological intervention tested | 0 (0%) | 2 (40%) | 11 (100%) |
| Non-sleep health outcomes measured | 0 (0%) | 2 (40%) | 3 (27%) |
