## Supplement 5 - instruments used to measure sleep for "Sleep interventions for adults admitted to psychiatric inpatient settings: a systematic scoping review"

**Supplement 5: Instrument used to measure sleep in the psychiatric inpatient setting**

| Perspective | Reporter | Instrument | No. of studies |
| --- | --- | --- | --- |
| Objective | Technology | PSG (86) | 1 |
|  |  | Actigraphy (52, 63) | 2 |
| Non-validated subjective | Staff | Nurse-led sleep chart/observations  (54, 57, 58, 61) | 4 |
|  | Patient | Unvalidated structured sleep questions (48, 58) | 2 |
|  |  | Sleep diary (46, 47) | 2 |
| Validated subjective | Patient | Pittsburgh sleep quality index (PSQI) (45, 46, 60, 63, 65) | 5 |
|  |  | Insomnia severity index (ISI) (50, 55, 62) | 3 |
|  |  | Richards-Campbell sleep questionnaire (RCSQ)(49, 87) | 2 |
|  |  | Daytime sleepiness scale (DSS)(47) | 1 |
|  |  | Dysfunctional beliefs and attitudes about sleep (DBAS) (65) | 1 |
|  |  | Epworth sleepiness scale (ESS)(46) | 1 |
|  |  | Fragebogen zur erfassung allgemeiner persönlichkeitsmerkmale schlafgestörter (FEPSIII) (50) | 1 |
|  |  | German sleep questionnaire A (SF-A)(88) | 1 |
|  |  | German sleep questionnaire B (SF-B)(88) | 1 |
|  |  | Nocturnal sleep onset scale (NSOS) (47) | 1 |
|  |  | Nursing outcome classification (NOC)(89) | 1 |
|  |  | Oviedo sleep questionnaire (OSQ)(89) | 1 |
|  |  | Pre-sleep arousal scale (PSAS) (65) | 1 |
|  |  | Sleep hygiene practice scale (SHPS)(65) | 1 |
|  |  | Stanford sleepiness scale (SSS) (46) | 1 |
